# Choroid plexus volume is enlarged in clinically isolated syndrome patients with optic neuritis

**DOI:** 10.1101/2022.08.23.22279105

**Authors:** Samuel Klistorner, Anneke Van der Walt, Michael H Barnett, Helmut Butzkueven, Scott Kolbe, John Parratt, Con Yiannikas, Alexander Klistorner

## Abstract

**Objectives:** People with Multiple Sclerosis (MS) have a larger choroid plexus (CP) volume than healthy controls. We investigated CP volume in early MS by quantitatively assessing brain MRI scans in patients presenting with optic neuritis (ON) as a clinically isolated syndrome (CIS), compared to a cohort with established Relapsing Remitting Multiple Sclerosis (RRMS) and healthy controls.

**Methods:** Pre- and post-gadolinium 3D-T1, 3D FLAIR and diffusion-weighted images were acquired from 44 CIS ON patients at baseline, 1, 3, 6 and 12 months after the onset of ON. Fifty RRMS patients and 50 healthy controls were also included for comparison.

**Results:** ANOVA revealed significantly larger CP volumes in both ON CIS and RRMS groups compared to healthy controls (p<0.001 for both), but no difference between ON CIS and RRMS patients (p=0.9)

Twenty-three ON CIS patients who converted to CDMS during 10 years of follow-up demonstrated CP volume similar to RRMS patients, but significantly larger compared to healthy controls (p<0.001). Increased CP volume was identified even in a sub-group of patients without MS-like lesions at baseline (p<0.001).

A significant (∼6%) transient increase of CP volume was observed following a new bout of inflammation, which, however, returned to pre-inflammatory state few months later. CP volume was not associated with the severity of acute inflammation of the optic nerve or long-term optic nerve axonal loss, not with brain lesion load or severity of tissue damage within lesions.

**Interpretation:** Our data demonstrate that enlarged CP can be observe very early in a disease, transiently reacts to acute inflammation, but not associated with the degree of tissue destruction.

## Introduction

The choroid plexus (CP), located within the brain’s ventricular system, principally produces cerebrospinal fluid (CSF) (Hofman and Chen 2016). It also plays an important role in the coordination of immune surveillance within the central nervous system (CNS), by regulating entry of immune cells and specific molecules into the brain parenchyma. Specifically, the CP provides a gateway for antigen trafficking between the CSF and the vascular circulation (Vercellino et al. 2008) (Monaco et al. 2020) (Silva-Vargas et al. 2016). Therefore, in contrast to the blood-brain barrier (BBB) and the blood-leptomeningeal barrier (BLMB), the blood-CSF barrier (BCSFB) functions as a controlled gate for immunosurveillance, and has a potential role in broader neuroimmune communication (Ayub, Jin, and Bae 2021)(Engelhardt, Wolburg-Buchholz, and Wolburg 2001).

Several recent reports have demonstrated that people with Multiple Sclerosis (MS) have a larger CP volume than healthy controls. Furthermore, the volume of the CP can be correlated to traditional measures of disease activity, including relapse rate, the number of gadolinium-enhancing MRI lesions and the expansion of chronic T2/Flair lesions (Fleischer et al. 2021) (V. A. Ricigliano et al. 2021) (Manouchehri and Stüve 2021) (S. Klistorner et al. 2022).

However, the temporal evolution of CP enlargement in MS is unknown. Therefore, to evaluate the CP volume in early MS, we quantitatively assessed brain MRI scans in patients presenting with optic neuritis (ON) as a clinically isolated syndrome (CIS), compared to a cohort with established Relapsing Remitting Multiple Sclerosis (RRMS) and age and sex-matched healthy controls.

## Methods

The study was approved by the University of Sydney, Macquarie University and Royal Melbourne Hospital Human Research Ethics Committees and followed the tenets of the Declaration of Helsinki. Written informed consent was obtained from all participants.

### Subjects

Data from forty-four patients enrolled in an ON study conducted at the Royal Melbourne Hospital between 2008-2010 were analysed retrospectively. All patients presented with acute unilateral ON diagnosed by identical clinical criteria used in the Optic Neuritis Treatment Trial (ONSG 1991). Participants were tested at baseline (i.e. within 48 hours of presentation), 1, 3, 6 and 12 months after the onset of ON.

For comparison, we included fifty consecutive patients with established RRMS, defined according to the revised McDonald 2010 criteria(Polman et al. 2011) (who are currently enrolled in the MADMS study) and 50 healthy controls. All groups had similar age and sex composition (see Table 1).

**Table 1.** Demographic data.

|  | ON CIS patients | RRMS patients | Healthy controls |
| --- | --- | --- | --- |
| N | 44 | 50 | 50 |
| Age, years (Mean±SD) | 33.9 ± 8.9 | 38.2 ± 9.3 | 36.0 ± 7.4 |
| F/M ratio | 2.3 | 2.1 | 2.3 |
| Disease duration, years (Mean±SD) | - | 5.4 ± 4.2 | - |
| EDSS at baseline (median (range)) | 3 (0-4) | 1 (0-4) | - |
| EDSS at 10 years follow-up (median (range)) | 0 (0-2) | - | - |
| Brain lesion volume, mm<sup>2</sup> (Mean±SD) | 1892 ± 4880 | 5769 ± 5229 | - |

### MRI protocol and analysis

CIS participants and fifteen healthy controls were imaged using a Siemens 3T Trio MRI system with a 32-channel receiver head coil. RRMS patients and 35 healthy controls were scanned using a 3T GE Discovery MR750 scanner (GE Medical Systems, Milwaukee, WI).

Specific acquisition parameters and MRI image processing are presented in Supplementary material.

CP within the lateral ventricles was semi-automatically segmented on T1 images using JIM 9 software (Xinapse Systems, Essex, UK) by a trained analyst (AK). The analyst, blinded to both clinical and lesion data, ensured that the correct contour was selected by the software and performed manual adjustments where needed. SienaX scaling was used to correct for inter-subject variability of head size.

The degree of tissue damage in chronic lesions was assessed by Mean Diffusivity (MD), as previously described (Castriota-scanderbeg et al. 2003) (A. Klistorner et al. 2018).

### Optical Coherence Tomography (OCT)

OCT was performed using an OCT-3 scanner (Stratus™, software version 3.0, Carl ZeissMeditec Inc.) using the Fast RNFL protocol consisting of three circular 3.4 mm diameter scans centred on the optic disc. A signal strength of seven or more was deemed acceptable. RNFL thickness was measured at 12 months for the affected and unaffected eyes. The difference in RNFL thinning between affected and unaffected eyes was used to minimise inter-subject variability.

### Multi-focal Visual Evoked Potentials (mfVEP)

The mfVEP was performed using the Accumap™ (ObjectiVision, software: Opera, Sydney, Australia) with a previously described testing procedure [<u>7</u>] that entailed recordings from 58 sectors of the visual field using four electrodes placed over the inion on the rear of the skull. Eyes were individually stimulated for 10 to 12 minutes until a sufficient signal-to-noise ratio (SNR) was reached. Mean latency was calculated per eye as a whole. The difference in mfVEP latency between affected and unaffected eyes was used to minimise inter-subject variability (A. Klistorner et al. 2008).

## Results

Demographic data are presented in Table 1. There were no significant differences in age or sex among the ON CIS, RRMS and healthy control cohorts.

### Comparison between ON CIS, clinically definite MS and healthy controls

No significant difference was observed between healthy controls imaged on Siemens and GE MRI scanners (1680 vs 1740, p=05). Therefore, data for healthy controls were combined.

There was a significant difference in CP volume between the three study groups (p<0.001, one-way ANOVA). Post-hoc analysis revealed significantly larger CP volumes in both ON CIS (2260 ± 534 mm^3^) and RRMS (2240 ± 659mm^3^) groups compared to healthy controls (1735 ± 338mm^3^, p<0.001 for both). There was no difference in CP volume between ON CIS and RRMS patients (p=0.9) (Fig.1).

**Fig. 1.**
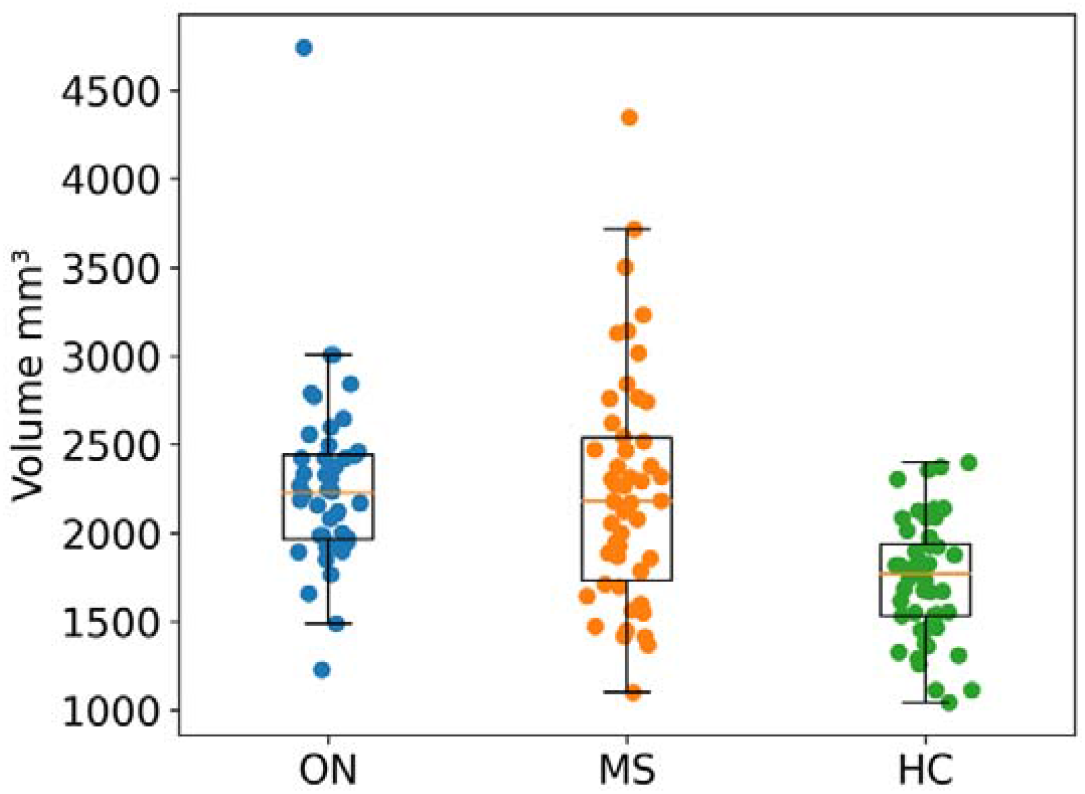
Normalized CP volume in CIS ON patients, RRMS patients and healthy controls.

Twenty-three ON CIS patients converted to clinically definite MS (CDMS) within ten years from the time of acute ON, while 21 ON CIS patients did not (diagram in Fig. 2).

**Fig. 2.**
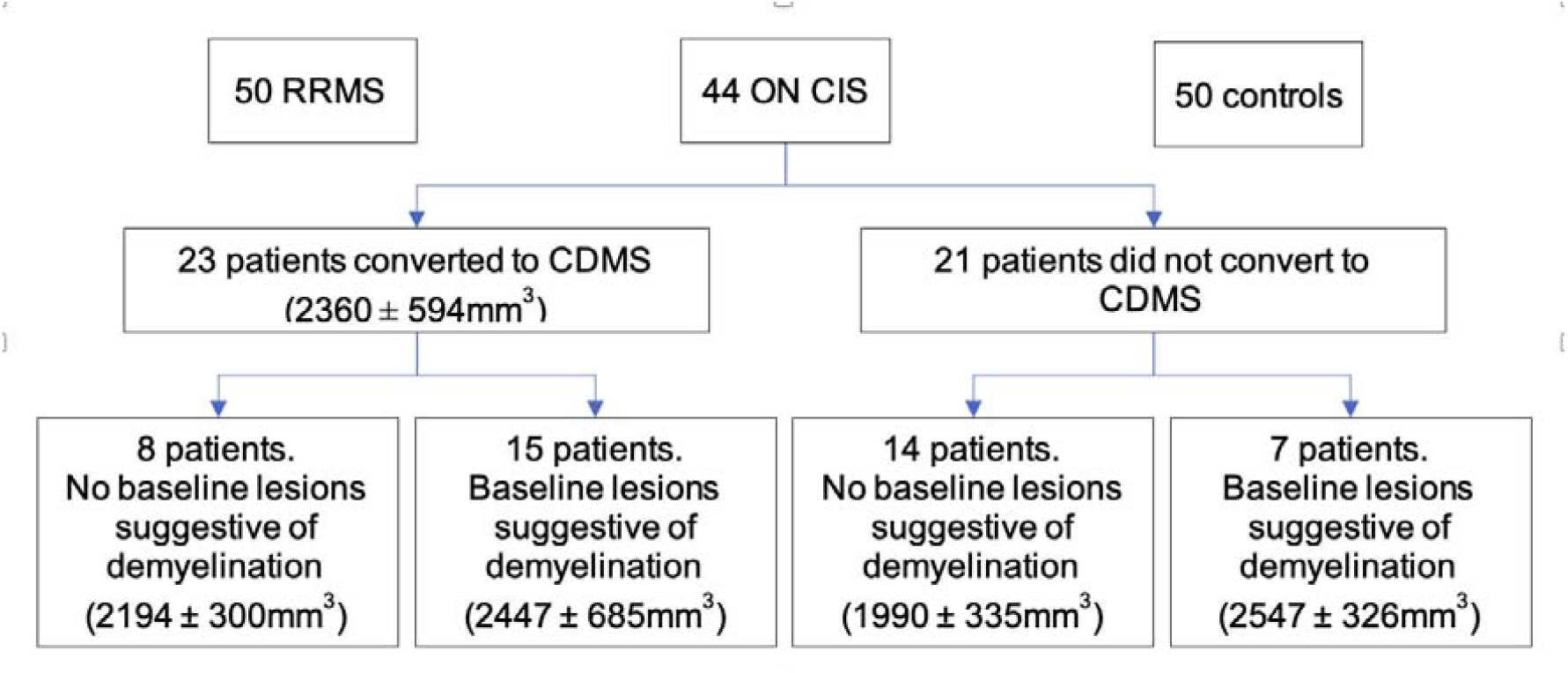
Diagram showing study groups.

### Choroid plexus volume is increased at the earliest stages of multiple sclerosis

Twenty-three ON CIS patients who converted to CDMS during 10 years of follow-up demonstrated CP volume similar to RRMS patients, but significantly larger compared to healthy controls (2360 ± 594mm^3^ vs 1735 ± 338mm^3,^ p<0.001).

Fifteen of those 23 patients demonstrated brain lesions suggestive of demyelinating disease at the time of ON presentation (average lesion volume: 1892 ± 4880 mm3). In six of those patients, at least one lesion was Gd-enhancing. Sub-analysis of CP in this group of 15 patients revealed significantly higher volume compared to healthy controls (2447 ± 685mm^3^ vs 1735 ± 338mm^3^, p<0.001). The remaining 8/15 patients were lesion-free at the time of ON presentation, suggesting that acute ON, and not preceding subclinical lesional activity, was the initial MS event. Sub-analysis of CP in this group also revealed significantly larger volume compared to normal controls (2194 ± 300mm^3^ vs 1735 ± 338mm^3^, p<0.001). There was no, however, significant difference in CP volume between two sub-groups of CIS patients (p=0.3).

In the 21/44 ON CIS patients who did not convert to CDMS during follow-up period, the CP was also significantly enlarged (p<0.001) compared to healthy controls. In this cohort, lesions suggestive of demyelination were observed on the baseline MRI in 7/21 patients. Compared to healthy controls, both patients with and without baseline brain demyelinating lesions demonstrated considerable enlargement of the CP (1990 ± 335mm^3^vs 1735 ± 338mm^3^, p=0.02 and 2547 ± 326mm^3^ vs 1735 ± 338mm^3^, p<0.001 respectively). CP enlargement, however, was significantly more pronounced in the former group (p=0.003).

Further analysis was restricted to the 23 patients who converted to CDMS during the ten years follow-up period.

### Temporal evolution of choroid plexus enlargement in CIS patients who converted to multiple sclerosis

To assess the effect of disease stage on CP enlargement, we estimated CP volume at one month and 12 months post ON in patients who ultimately converted to CDMS with no brain demyelinating lesions (n=8) at baseline. There was no difference in CP volume between baseline and follow-up time points (average CP volumes 2194 ± 300mm^3^, 2213 ± 314mm^3^ and 2195± 330mm^3,^ respectively, ANOVA p>0.05).

To investigate the effect of new bouts of inflammation on plexus size, the change of plexus volume (as measured from the previous timepoint) in a sub-group of ON CIS patients who demonstrated new lesion activity (i.e. new T2 lesions > 3 mm in diameter) during the initial 12 months follow-up period was examined. We selected only those patients who developed new lesions at 3, 6 or 12 months after the onset of ON.; further, we confirmed that there were no lesion activity in the interval directly preceding the development of the new lesion (i.e. between 1 and 3 months, 3-6 months or 6-12 months respectively). These criteria were satisfied in 14/23 patients (Fig. 3).

**Fig. 3.**
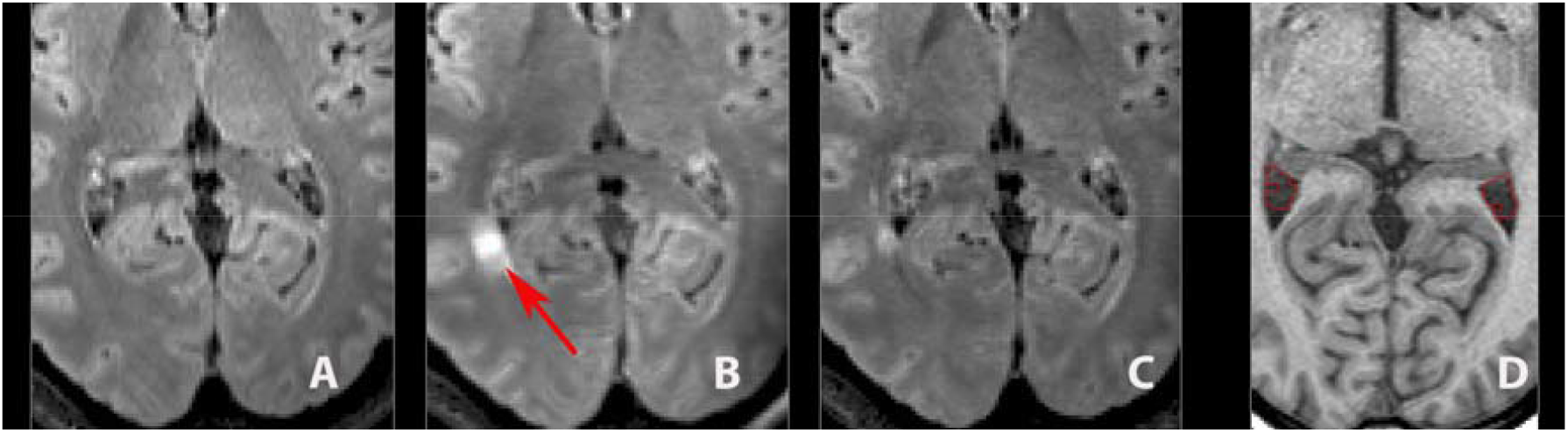
Example of patient with new lesion at 3 months (arrow). a. FLAIR image at 1 month (pre-lesion) b. FLAIR image at 3 month (acute lesion) c. FLAIR image at 6 month (post-lesion) d. T1 image with plexus masking

A significant increase in CP volume was observed in patients with new lesion activity (2670+/-730 vs 2833+/- 756 respectively, p<0.001 paired t-test, representing an average increase of 6.3+/-3.9%). In 10/14 cases examined, new lesions occurred at 3 or 6 months, providing an opportunity to examine changes in CP volume during the post-inflammatory period (i.e. at 6 or 12 months). A significant increase of CP volume was observed within 2-3 months of a new bout of inflammation evident on MRI at 3 or 6 months (2431+/-468 vs 2593+/-496 for pre-and inflammatory stages respectively, representing an increase of 6.7+/- 4%, p<0.001). CP volume returned almost to its pre-inflammatory state at 6 or 12 months respectively (2477+/-477 vs 2431+/-468 for pre-and post-inflammatory stages respectively, or difference of 1.4+/-4%, p=0.1).

**Fig. 3.**
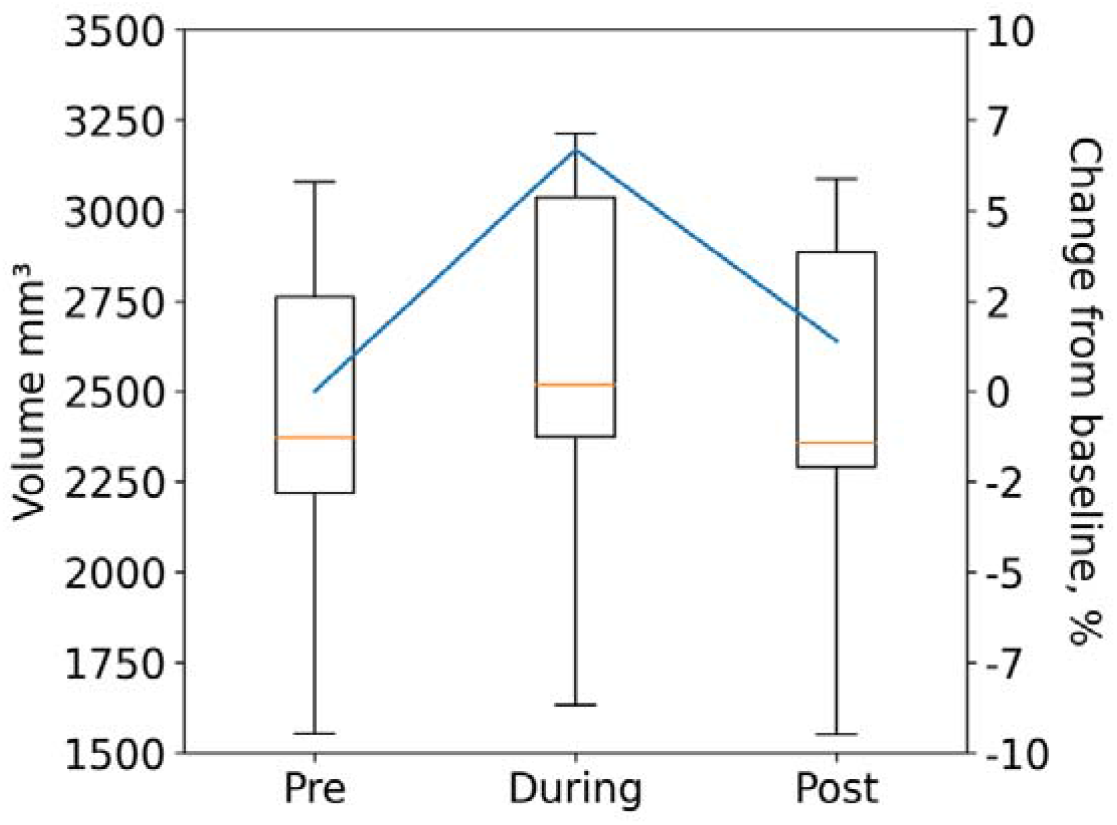
Box-plot of CP volume before (“Pre”), during and after (“Post”) new lesional activity.

### CP volume is not associated with the severity of acute inflammation of the optic nerve

Measurement of the optic nerve lesion length during acute ON was performed in all 23 ON CIS patients who converted to CDMS. The average ON lesion length was 10.7 ± 5.8mm. Due to a high incidence of conduction block at the time of acute ON, analysable mfVEP waveforms were obtained only in 15/23 patients at baseline MRI testing. The average mfVEP latency delay in this group, in the ON eye compared to the fellow eye, was 11.4 ± 9.7 msec.

Both measures of acute inflammation (lesion length and mfVEP latency delay) demonstrated a wide spectrum of values (ON lesion length: 0-21.5 mm; inter-eye mfVEP latency asymmetry: 2-22 msec), indicating a variable degree of acute demyelination in the optic nerve. However, while these metrics were highly intracorrelated (r=0.78, p<0.001), neither were associated with CP volume.

### CP volume is not associated with long-term optic nerve axonal loss

Measurement of RNFL thickness at 12 months post-acute ON was available 21/23 ON CIS patients who converted to CDMS; and revealed a marked loss of the RNFL in affected (ON) eyes compared to fellow eyes (inter-eye asymmetry 15.0 ± 11.2 microns), indicating considerable loss of RGC axons caused by optic nerve inflammation. No correlation was observed between baseline CP volume and relative RNFL loss in ON eyes.

### CP volume and brain inflammatory load in ON CIS patients

Data analysis from 23 ON CIS patients who converted to CDMS showed no association of CP volume with total brain T2 lesion volume, number of brain T2 lesions or average brain T2 lesion size.

Average (patient-wise) MD in the core of chronic lesions varied from values close to the MD typically observed in NAWM (0.87) to highly abnormal values (1.4), indicating a broad spectrum of lesional tissue damage. However, there was no correlation between average MD in the core of chronic lesions and CP volume (Gadolinium-enhancing lesions were excluded from diffusivity analysis to minimise the confounding effect of oedema related to acute inflammation).

## Discussion

The choroid plexus, a key gateway between the systemic circulation and the CNS, has recently attracted the attention of MS researchers. Indeed, several recent studies have identified a relationship between CP volume and the degree of acute and chronic inflammation in brain tissue.(Fleischer et al. 2021) (Manouchehri and Stüve 2021) (V. A. Ricigliano et al. 2021) (S. Klistorner et al. 2022). We recently reported that the volume of CP predicts subsequent expansion of chronic periventricular MS lesions and is associated with tissue damage within and outside of chronic lesions.

In the current study, we examined the size of CP in a cohort of CIS patients presenting with acute ON, and investigated the relationship between CP volume and various measures of inflammatory demyelination and axonal loss both in the visual system and brain.

Our principal finding was an increased CP volume in ON CIS patients, suggesting that the pathomechanisms that underpin this phenomenon is present at the earliest stages of MS. Enlargement of the CP was even observed in a subgroup of patients who did not display brain lesions suggestive of demyelinating disease at the time of ON presentation but later converted to CDMS. The absence of lesions compatible with demyelination suggests that ON may be the initial inflammatory event in this sub-group. Our findings, therefore, suggest that CP enlargement may be relevant to the pathogenesis of further inflammatory demyelination that results in clinically definite MS. CP enlargement, therefore, can precede the diagnosis of MS, even when this is based on modern diagnostic criteria that incorporate MRI characteristics that represent ongoing inflammatory activity in space and time. This is consistent with a recently reported finding of enlarged CP in a cohort of RIS patients (V. A. Ricigliano et al. 2021) (V. Ricigliano, Louapre, and Poirion 2021). In another study that demonstrated enlargement of the CP, a significant proportion of participants was also represented by CIS patients (Fleischer et al. 2021).

We also observed a small but significant transient enlargement of the CP following new bouts of inflammation (as determined by newly appearing T2 brain lesions). Since new lesions appeared within 2-3 months preceding MRI examination (i.e. were absent at a previous scan), it is reasonable to assume that inflammation still remained active at the time of the MRI scanning following acute lesions but is likely to subside by the next scan (3-6 months later). This finding agrees with recently published experimental data describing transient enlargement of the CP in both cuprizone and experimental allergic encephalomyelitis (EAE) models of inflammatory demyelination (Fleischer et al. 2021). CP volume in our study returned to its pre-inflammatory state within a few months, recapitulating the experimental data. Several potential explanations for transient CP enlargement, such as increased migration of immune and glial cells into the brain parenchyma, oxidative CP injury, increased permeability of CP epithelial cells or inflammation of the CP itself, have been suggested (Fleischer et al. 2021) (Vercellino et al. 2008) (Rodríguez-Lorenzo et al. 2020). A marked reduction in neuro-inflammation may explain (at least to some extent) the recent observation that pharmacological modulation of the blood-CSF barrier by natalizumab, an efficacious anti-inflammatory therapy(Samjoo et al. 2021), appears to prevent CP enlargement, compared to untreated patients and patients receiving dimethyl fumarate (Fleischer et al. 2021).

However, the mechanisms underlying structural changes that lead to permanently increased CP volume in early MS remain largely unknown. Intriguingly, enlargement of the CP has also been reported in other neurological conditions which exhibit some degree of neuroinflammation, including acute stroke(Egorova et al. 2019), psychosis(Lizano et al. 2019), schizophrenia(Y. F. Zhou et al. 2020), depression(Althubaity et al. 2022), Alzheimer disease(Choi et al. 2022) and complex regional pain syndrome(G. Zhou et al. 2015). Therefore, enlargement of the CP is not MS-specific but instead is likely to be associated with general increase of neuroinflammation or neuronal/axonal degeneration (Althubaity et al. 2022) (Choi et al. 2022). Indeed, functional dysregulation of the CP may reflect yet unknown common underlying mechanisms in the pathophysiology of neurodegenerative diseases (Schwartz and Cahalon 2022).

Furthermore, it has been suggested that enlargement of the CP may occur before the development of pathological changes in the CNS and may even reflect an inherited or acquired susceptibility to neuroinflammation(Egorova et al. 2019) (Althubaity et al. 2022) (Y. F. Zhou et al. 2020). For example, enlargement of the CP has been reported in stroke patients in the early, post-acute period. CP enlargement did not respect stroke laterality and remained stable during 12 months of follow-up, prompting the authors to raise the possibility that this phenomenon predated the stroke and may be “associated with increased vascular burden and neuroinflammation, which may also increase the likelihood of stroke” (Egorova et al. 2019). In patients with recently diagnosed schizophrenia, significantly larger CP volumes, compared with healthy controls were observed, leading the authors to speculate that “it is possible that this (CP enlargement) may occur prior to the first episode of schizophrenia” or even be related to abnormal adult neurogenesis (Y. F. Zhou et al. 2020). Moreover, a similar study of patients with psychosis not only demonstrated increased CP volume across the disease spectrum but also reported significant heritability of this observation by examining first-degree relatives(Lizano et al. 2019).

The finding of early CP enlargement in other neurological conditions is in line with our observation that increased CP volume can be observed at the initial stages of the disease and even at the time of first MS attack. While a smaller CP volume was found in CDMS patients without baseline lesions compared to patients with demyelinating brain lesions at baseline (i.e. patients likely to have a longer disease duration), this difference was not statistically significant. Furthermore, the stability of longitudinal CP data within the first 12 months of post-ON also does not support the scenario of gradual plexus enlargement. We understand, however, that this may be affected by small group size and limited duration of follow up.

CP volume in ON CIS patients was not associated with severity of acute inflammation of the optic nerve, as measured by MRI-defined lesion length and extent of acute demyelination determined by latency delay of the mfVEP. There was also no relationship between CP size and axonal loss resulting from optic neuritis (as measured by relative thinning of RNFL). Likewise, CP volume was not related to volumetric characteristics of brain lesions or the degree of lesional damage measured by tissue rarefication (MD change). Analogously, a similar lack of relationship between stroke size and CP volumes has recently been reported in stroke patients (Egorova et al. 2019). Caution, however, must be taken in interpreting this finding since the starting point of the disease may precede the first clinical attack by several years.

Although patients who did not convert to CDMS in our study also demonstrated increased CP volume, this was primarily driven by the subjects with demyelinating brain lesions on baseline imaging, a cohort that remains at high risk of developing MS even 10 years after acute optic neuritis (ONSG 2008). Our observation may therefore reflect the mixed (MS vs non-MS) nature of the non-converter group.

Our study has several limitations. The sample size, particularly for sub-analyses of patients with and without baseline brain lesions, was small and potentially impacts the robustness of our conclusions. Since the source data for our study was collected more than ten years ago, a first-generation OCT device was used to analyse RNFL thickness. However, the effect of ON on the axonal loss of retinal ganglion cells is profound, and the accuracy of time-domain OCT, therefore sufficient for this purpose. To further reduce measurement variability, relative inter-eye asymmetry, rather than absolute RNFL thickness, was used.

### Conclusion

The CP is enlarged in ON CIS patients, even when ON is the only discernible MS pathology on MRI. In addition, there is a transient enlargement of the CP in association with the appearance of new brain lesions. However, the pathophysiology of CP enlargement in MS is currently unknown and may include several mechanisms. Similar to other neurodegenerative diseases, it remains to be seen whether enlargement of the CP is a cause or an effect of CNS neuroinflammation(Gião et al. 2022) (Choi et al. 2022) (Y. F. Zhou et al. 2020).

## Data Availability

 All data produced in the present study are available upon reasonable request to the authors

## Supplementary material

### Siemens 3T Trio protocol

a. Pre- and post-contrast (gadolinium) 3D T1-weighted volumetric sequence (TR = 1900⍰ms, TE = 2.631ms, TI = 900⍰ms, flip angle = 9°, voxel dimensions = 0.8 × 0.8 × 0.8⍰mm^3^)
b. 3D T2-weighted FLAIR sequence (TR = 50001ms, TE = 436⍰ms, TI = 2100⍰ms, flip angle = 120°, voxel dimensions = 0.9 × 0.9 × 0.91mm^3^, GRAPPA parallel speed factor = 2, and acquisition time = 7⍰m 37⍰s).
c. ,echo-planar imaging diffusion-weighted sequence (TR = 8200⍰ms; TE = 89⍰ms; voxel dimensions = 2.0 × 2.0 × 2.0⍰mm^3^; 60 directions b = 1200⍰s/mm^2^).

### 3T GE Discovery MR750 protocol

a. Pre- and post-contrast (gadolinium) Sagittal 3D T1 (GE BRAVO sequence, duration 4 min each, FOV 256mm, Slice thickness 1mm, TE = 2.7ms, TR = 7.2ms, Flip angle 12°, voxel dimensions = 1 × 1 × 1⍰mm^3^).
b. GE CUBE T2 FLAIR sequence (TR = 8000ms TE = 163ms, Flip angle 90°, voxel dimensions = 0.9 × 0.9 × 1.2⍰mm^3^ and acquisition time = 6 min).
c. Echo-Planar Imaging diffusion-weighted sequence (TR = 8325; TE = 86 ms, voxel dimensions = 2.0 × 2.0 × 2.0⍰mm^3,^ duration 9 min, 64-directions, b = 1000 s/mm^2^).

### MRI image pre-processing

The baseline T1-weighted imaging was realigned to Anterior and Posterior Commissure (AC-PC) orientation. Using FLIRT (FSL, FMRIB Software Library), follow-up T1 images were co-registered to initial (month 0) AC-PC space by applying transformation matrices derived from linear co-registration between baseline AC-PC aligned brain and follow-up native T1 brain images. In parallel, diffusion MRI was corrected for motion and eddy-current distortion in FSL, then EPI susceptibility distortion was minimized by applying deformation maps generated from nonlinear co-registration between DWI b0 brain images and T1-weighted images at each time-point using ANTS (Advanced Normalization Tools). Subsequently, tensor reconstruction was performed in MRtrix3. Tensor and FLAIR images were then linearly co-registered to corresponding T1 AC-PC images at each timepoint.

### DTI data processing

Diffusion weighted MRI data (dMRI) were pre-processed using tools provided by the software suites MRtrix3 [1], FSL [2], and ANTs [3, 4]. Specifically, dMRI data were first denoised [5], then potential Gibbs-ringing artefacts were removed [6]. The dMRI data were then corrected for bias field inhomogeneities using the ANTs N4 algorithm [4]. a dMRI brain mask was then estimated using BET [7] and used as an input, alongside the dMRI data, to eddy [8] in order to correct for subject movement in the acquisition. Finally, phase distortion correction was applied using a non-linear registration method outlined below.

To correct for phase distortion within the dMRI data, first the brain was segmented from the corresponding T1w dataset, and a single B0 volume was extracted from the dMRI dataset. The T1w brain was then used as a mask to invert the contrast of the T1w image. A rigid-body registration was then performed on the inverted T1w image with the B0 volume as a target, which aligned the two images spatially. Non-linear registration was then performed using ANTs. The registration steps were comprised of a rigid body, then affine, and then SyN registration algorithm [3]. The transformations and warps calculated from the non-linear registration steps were then applied to the entire dMRI dataset to correct for phase distortion artefacts.

## References

Althubaity, Noha et al. 2022. “Choroid Plexus Enlargement Is Associated with Neuroinflammation and Reduction of Blood Brain Barrier Permeability in Depression.” NeuroImage: Clinical 33. http://creativecommons.org/licenses/by/4.0/ (April 10, 2022).

Ayub, Maria, Hee Kyung Jin, and Jae sung Bae. 2021. “The Blood Cerebrospinal Fluid Barrier Orchestrates Immunosurveillance, Immunoprotection, and Immunopathology in the Central Nervous System.” BMB Reports 54(4): 196–202.

Castriota-scanderbeg, Alessandro et al. 2003. “Coefficient Dav Is More Sensitive Than Fractional Anisotropy in Monitoring Progression of Irreversible Tissue Damage in Focal Nonactive Multiple Sclerosis Lesions.” AJNR Am J Neuroradiol 24(April): 663–70.

Choi, Jong Duck et al. 2022. “Choroid Plexus Volume and Permeability at Brain MRI within the Alzheimer Disease Clinical Spectrum.” Radiology (9).

Egorova, Natalia et al. 2019. “Choroid Plexus Volume after Stroke.” International Journal of Stroke 14(9): 923–30.

Engelhardt, Britta, Karen Wolburg-Buchholz, and Hartwig Wolburg. 2001. “Involvement of the Choroid Plexus in Central Nervous System Inflammation.” Microscopy Research and Technique 52(1): 112–29.

Fleischer, Vinzenz et al. 2021. “Translational Value of Choroid Plexus Imaging for Tracking Neuroinflammation in Mice and Humans.” Proceedings of the National Academy of Sciences of the United States of America 118(36): 1–12.

Gião, Tiago, Tiago Teixeira, Maria Rosário Almeida, and Isabel Cardoso. 2022. “Choroid Plexus in Alzheimer’s Disease—The Current State of Knowledge.” Biomedicines 10(2).

Hofman, Florence M, and Thomas C Chen. 2016. “Choroid Plexus: Structure and Function.” In THE CHOROID PLEXUS AND CEREBROSPINAL FLUID, eds. Josh Neman and Thomas C Chen. Amsterdam: Elsevier Inc., 29–40.

Klistorner, A. et al. 2008. “Axonal Loss and Myelin in Early ON Loss in Postacute Optic Neuritis.” Ann Neurol 64(3): 325–31. http://www.ncbi.nlm.nih.gov/pubmed/18825673.

Klistorner, A. et al. 2018. “Evidence of Progressive Tissue Loss in the Core of Chronic MS Lesions: A Longitudinal DTI Study.” NeuroImage: Clinical 17: 1028–35.

Klistorner, Samuel et al. 2022. “Choroid Plexus Volume Predicts Expansion of Chronic Lesions and Associated Brain Atrophy in Multiple Sclerosis.” Annals of Clinical and Translational Neurology ccepted.

Lizano, Paulo et al. 2019. “Association of Choroid Plexus Enlargement with Cognitive, Inflammatory, and Structural Phenotypes across the Psychosis Spectrum.” American Journal of Psychiatry 176(7): 564–72.

Manouchehri, Navid, and Olaf Stüve. 2021. “Choroid Plexus Volumetrics and Brain Inflammation in Multiple Sclerosis.” Proceedings of the National Academy of Sciences of the United States of America 118(40): 10–12.

Mihaljevic, Sandra, Alena Michalicova, Mangesh Bhide, and Andrej Kovac. 2021. “Pathophysiology of the Choroid Plexus in Brain Diseases.” Gen. Physiol. Biophys. 40: 443–62.

Monaco, Salvatore, Richard Nicholas, Richard Reynolds, and Roberta Magliozzi. 2020. “Intrathecal Inflammation in Progressive Multiple Sclerosis.” International Journal of Molecular Sciences 21(21): 1–11.

ONSG. 1991. “The Clinical Profile of Optic Neuritis: Experience of the Optic Neuritis Treatment Trial.” Arch Ophthalmol 109: 1673–78.

ONSG. 2008. “Multiple Sclerosis Risk after Optic Neuritis: Final Optic Neuritis Treatment Trial Follow-Up.” Arch neurol 65: 727–32.

Polman, Chris H et al. 2011. “Diagnostic Criteria for Multiple Sclerosis: 2010 Revisions to the McDonald Criteria.” Annals of neurology 69(2): 292–302. http://www.pubmedcentral.nih.gov/articlerender.fcgi?artid=3084507&tool=pmcentrez&rendertype=abstract (July 10, 2014).

Ricigliano, VAG, Céline Louapre, and Émilie Poirion. 2021. “An Imaging Signature in Choroid Plexuses in Pre-Symptomatic Multiple Sclerosis.” Presented at Annual Meeting of the European Charcot Foundation Poster.

Ricigliano, Vito AG et al. 2021. “Choroid Plexus Enlargement in Inflammatory Multiple Sclerosis.” Radiology 301: 166–77.

Rodríguez-Lorenzo, Sabela et al. 2020. “Inflammation of the Choroid Plexus in Progressive Multiple Sclerosis: Accumulation of Granulocytes and T Cells.” Acta Neuropathologica Communications 8(1): 1–13.

Samjoo, Imtiaz A. et al. 2021. “Efficacy Classification of Modern Therapies in Multiple Sclerosis.” Journal of Comparative Effectiveness Research 10(6): 495–507.

Schwartz, Michal, and Liora Cahalon. 2022. “The Vicious Cycle Governing the Brain– Immune System Relationship in Neurodegenerative Diseases.” Current Opinion in Immunology 76: 102182. https://doi.org/10.1016/j.coi.2022.102182.

Silva-Vargas, Violeta et al. 2016. “Age-Dependent Niche Signals from the Choroid Plexus Regulate Adult Neural Stem Cells.” Cell Stem Cell 19(5): 643–52. http://dx.doi.org/10.1016/j.stem.2016.06.013.

Vercellino, Marco et al. 2008. “Involvement of the Choroid Plexus in Multiple Sclerosis Autoimmune Inflammation: A Neuropathological Study.” Journal of Neuroimmunology 199(1–2): 133–41.

Zhou, Guangyu et al. 2015. “Enlargement of Choroid Plexus in Complex Regional Pain Syndrome.” Scientific Reports 5: 1–5. http://dx.doi.org/10.1038/srep14329.

Zhou, Yan Fang et al. 2020. “Choroid Plexus Enlargement and Allostatic Load in Schizophrenia.” Schizophrenia Bulletin 46(3): 722–31.

